# Rapid detection of neutralizing antibodies to SARS-CoV-2 variants in post-vaccination sera

**DOI:** 10.1101/2021.05.06.21256788

**Authors:** Kei Miyakawa, Sundararaj Stanleyraj Jeremiah, Hideaki Kato, Yutaro Yamaoka, Hirofumi Go, Takeharu Yamanaka, Akihide Ryo

**Author notes:** **Corresponding author and address:** Akihide Ryo, M.D., Ph.D, Department of Microbiology and Molecular Biodefense Research, School of Medicine, Yokohama City University, 3-9, Fukuura, Kanazawa, Yokohama, Japan, 236-0004.

## Abstract

The uncontrolled spread of the COVID-19 pandemic has led to the emergence of different SARS-CoV-2 variants across the globe. The ongoing global vaccination strategy to curtail the COVID-19 juggernaut, is threatened by the rapidly spreading Variants of Concern (VOC) and other regional mutants, which are less responsive to neutralization by infection or vaccine derived antibodies. We have previously developed the hiVNT system which detects SARS-CoV-2 neutralizing antibodies in sera in less than three hours. In this study, we modify the hiVNT for rapid qualitative screening of neutralizing antibodies (nAb) to multiple variants of concern (VOC) of SARS-CoV-2, and assess the neutralizing efficacy of the BNT162b2 mRNA vaccine on seven epidemiologically relevant SARS-CoV-2 variants. Here we show that the BNT162b2 mRNA vaccine can activate humoral immunity against the major SARS-CoV-2 mutants that are currently in circulation. Albeit a small sample size, we observed that one dose of vaccine was sufficient to elicit a protective humoral response in previously infected people. Using a panel of seven SARS-CoV-2 variants and a single prototype virus, our modified hiVNT would be useful for large-scale community wide testing to detect protective immunity that may confer vaccine/immune passport in the ongoing COVID-19 pandemic.

## Introduction

The uncontrolled spread of the COVID-19 pandemic has led to the emergence of different SARS-CoV-2 variants across the globe. The ongoing global vaccination strategy to curtail the COVID-19 juggernaut, is threatened by the rapidly spreading Variants of Concern (VOC) and other regional mutants, which are less responsive to neutralization by infection or vaccine derived antibodies ^1,2^.

We have previously developed the hiVNT (HiBiT-based virus-like particle neutralization test) system which detects SARS-CoV-2 neutralizing antibodies in sera in less than three hours ^3^. It uses lentivirus-based virus-like particles (VLP) incorporated with the NanoLuc fragment peptide, HiBiT. Viral entry into reporter cells that express LgBiT intracellularly, allows the viral HiBiT to fuse with the LgBiT to reconstitute whole NanoLuc luciferase which is readily detected by luminometer ^3,4^. The hiVNT assay is capable of high throughput and can be easily carried out in a low biosafety setting. We have modified the hiVNT for rapid qualitative screening of neutralizing antibodies (nAb) to multiple variants of concern (VOC) of SARS-CoV-2. In the current study, we assess the neutralizing efficacy of the BNT162b2 mRNA vaccine on a panel of seven epidemiologically relevant SARS-CoV-2 variants.

## Results

### Establishment of a modified hiVNT for qualitative detection of SARS-CoV-2 nAb

Using the spikes of the prototype virus (D614G) and seven variants (B.1.1.7, B.1.351, P.1, R.1, B.1.617, B.1.429, and B.1.526) (Figure 1A), we generated HiBiT-containing VLPs (hiVLP). If the serum nAbs neutralize the mutant hiVLPs, their entry into the reporter cells is prevented, reducing the luminescence emitted (Figure 1B). We optimized the conditions of the hiVNT using sera of COVID-19 patients (n=97) and healthy individuals (n=43) with pre-defined neutralization titers (NT50) by the pseudovirus-based neutralization assay. At a fixed serum dilution of one in twenty, the hiVNT assay could detect the nAb titers corresponding to NT50 of around 50 which is the positive cut-off in the standard pseudovirus neutralization assay. We then determined that a sample possessed neutralization activity if it showed more than 40% luminescence signal inhibition in the hiVNT assay (Figure 1C). The receiver operating characteristic (ROC) curve showed that this cut-off value of 40% can detect the presence of nAbs with a high degree of accuracy (Figure 1D). This modification of performing the hiVNT at a fixed dilution makes the procedure less laborious, reduces the turn-around-time while achieving performance similar to the standard neutralization assays.

**Figure 1.**
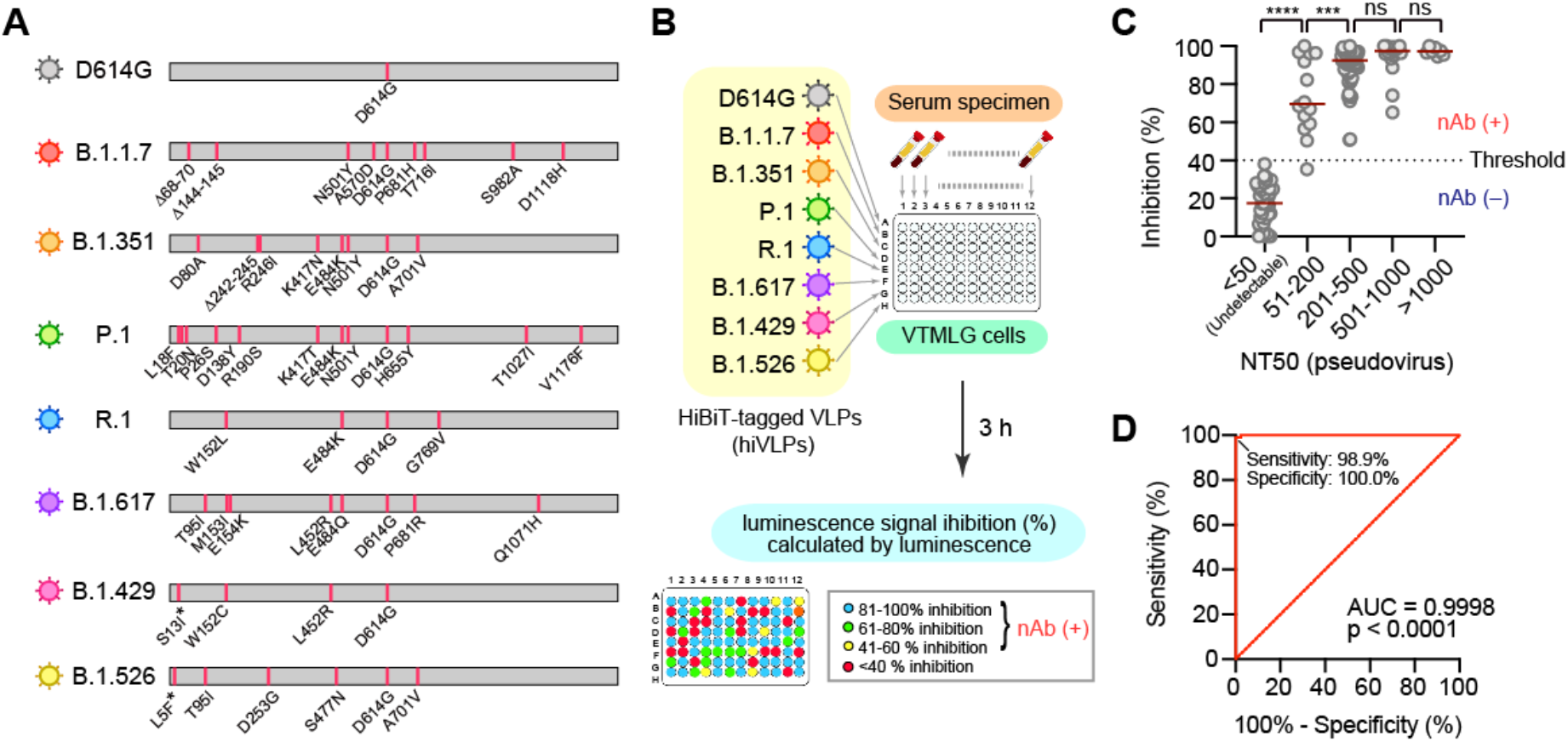
Modified hiVNT for qualitative detection of SARS-CoV-2 nAb. **(A)** Spike mutations of indicated SARS-CoV-2 variants used in this study. Since the N-terminus of the spike has been replaced with the CD5 signal sequence for incorporation into VLPs, the mutations marked with an asterisk are not applied in this study. **(B)** Eight strains of hiVLPs (HiBiT-Tagged VLPs) were added to VeroE6/TMPRSS2-LgBiT cells (VTMLG) in the presence of 5% of serum. Three hours later, cells were washed and measured luminescence to detect the luminescence signal inhibition of each serum. **(C)** Correlation of hiVNT with pseudovirus NT. Sera with the luminescence signal inhibition <40% can be deemed negative for neutralizing antibodies. ***p < 0.001, ****p < 0.0001, ns, not significant, two-tailed unpaired t-test. **(D)** ROC curve of hiVNT. Area under curve (AUC) is also shown. The cut-off value of 40 detects the presence of nAbs with a high degree of accuracy (Sensitivity 98.9%; Specificity 100%).

### Neutralizing activity of vaccinated sera against SARS-CoV-2 VOCs

The study samples were collected from otherwise healthy medical personnel who were administered with two doses of the vaccine three weeks apart. Serum of the vaccinees was collected at three instances: before the first dose, two weeks after first dose and one week after second dose. All sera collected prior first vaccine shot was checked for seropositivity using NP-IgG detection assay ^5^ to get two vaccine recipient groups: previously uninfected (n=105) and post-COVID-19 (n=6).

Among the previously uninfected vaccine recipients, 57.1% (60/105) showed a positive conversion for nAbs against the conventional strain (D614G) after the first dose. However, the nAbs against the mutant strains (except B.1.526) were much lower (16.2-39.0%) (Figure 2A, middle panel).

**Figure 2.**
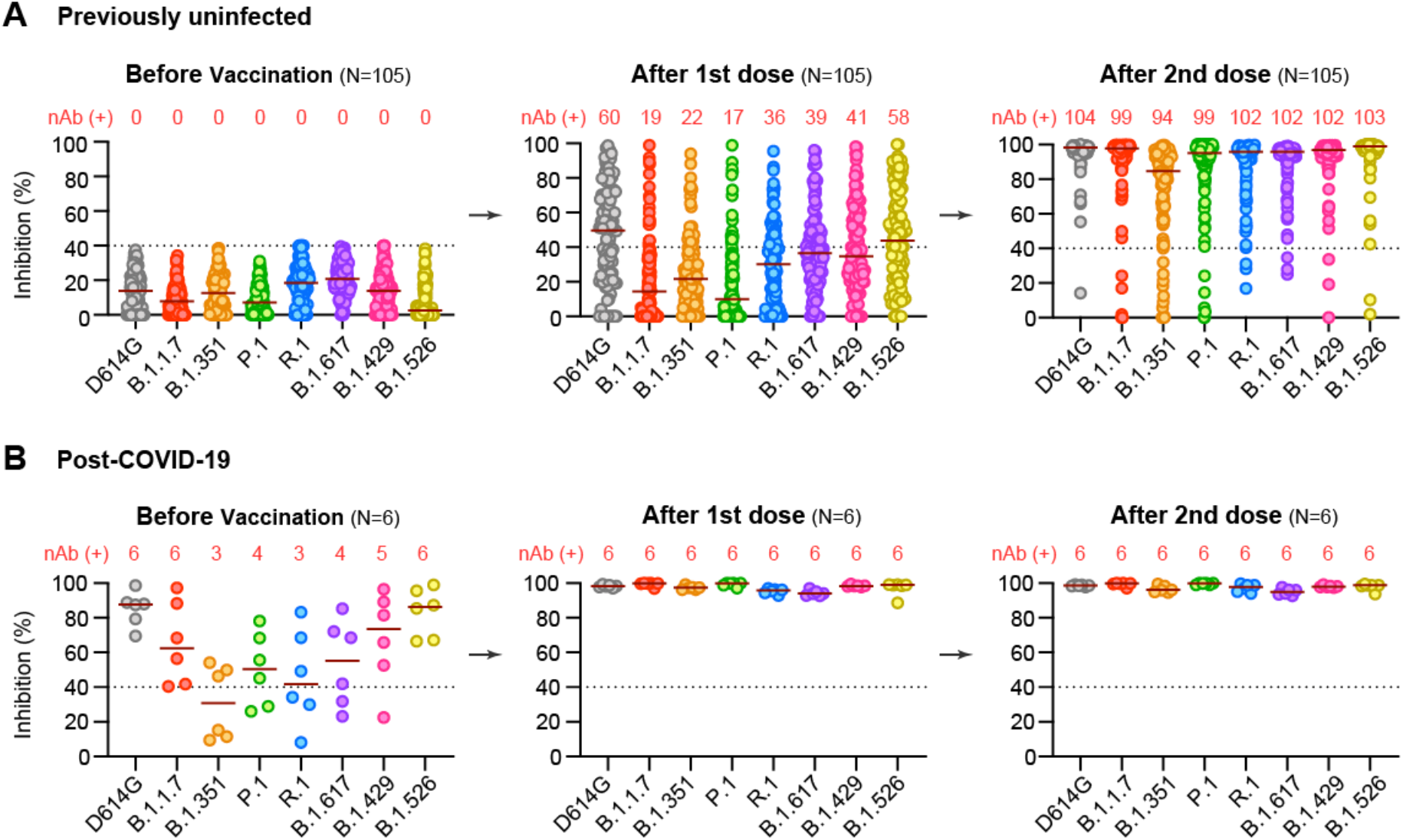
Positive conversion of nAb to SARS-CoV-2 variants in post-vaccination sera. **(A, B)** Positive conversion of nAb to SARS-CoV-2 variants in post-vaccination sera from previously uninfected (A) or post-COVID-19 (B). Numerals in red indicate the number of sera positive for nAb. Red horizontal bars represent median of luminescence signal inhibition.

A steady increase in nAbs was observed after the first dose, followed by the achievement of significant nAb levels against all mutants in most people after the second dose (Figure 2A, right panel). The vaccine derived nAbs possessed high median inhibition rates of above 94% for all variants except B.1.351 which was 89.5% (94/105). All the six individuals with past COVID-19 infection achieved significant levels of nAb after the first dose despite some possessing nAbs below threshold prior initial vaccination (Figure 2B).

## Discussion

Fitter mutants of SARS-CoV-2 of varying dissimilarity to the initial wild-type outbreak strain have emerged and disseminated rapidly. This has raised concerns about the efficacy of the currently available vaccines which rely on the antigenic determinants of the older virus to provide protective immunity. Among the currently studied mutants, those possessing the E484K mutation such as the B.1.351, P.1, and R.1 are notorious for escaping infection and vaccine derived nAbs ^2^.

To clear this doubt on vaccine escape, we developed an extensive mutant panel including the recently surging B.1.617 double mutant (E484Q and L452R) which is presumed to have higher propensity to escape nAbs. In this first report of vaccine efficacy assessment from Japan, we reveal that the BNT162b2 mRNA vaccine has adequate efficacy against the epidemiologically relevant SARS-CoV-2 variants. Vaccine escape was noted in all mutants but only in a minor fraction, which was higher for the B.1.351 mutant. Albeit a small sample size, we observed that one dose of vaccine was sufficient to elicit protective humoral response in previously infected people as documented by previous reports ^6^. Despite the presence of their corresponding nAbs in the serum, mutants can cause vaccine breakthrough infections, suggesting that the circulating nAbs might not offer protection at mucosal surfaces to prevent infection ^7^.

In the current study, we show that the BNT162b2 mRNA vaccine can activate humoral immunity against the major SARS-CoV-2 mutants that are currently in circulation. Our modified hiVNT possesses the advantages of being rapid, safe, concise and comprehensive over the conventional neutralization assays. However, this modified hiVNT assay is qualitative and cannot quantify the nAb titers at relatively higher levels that maybe relevant to their long-term persistence and duration of protection. Using a panel of seven SARS-CoV-2 variants and a single prototype virus, our modified hiVNT would be useful for large-scale community wide testing to detect protective immunity that may confer vaccine/immune passport in the ongoing COVID-19 pandemic. The selection pressure exerted by the current vaccines would cause the evolution of vaccine escape mutants, which must be monitored in the future and for which the hiVNT assay developed using the spikes of new variants could come in handy.

## Materials and Methods

### Ethics statement

This study was approved by Yokohama City University Certified Institutional Review Board (Reference No. B160800009, B200600115, B210300001), and the protocols used in the study were approved by the ethics committee. Written informed consent was obtained from all the participants.

### Production of hiVLP

HEK293 cells (ATCC #CRL-1573), VeroE6/TMPRSS2 cells (JCRB #1819), and VeroE6/TMPRSS2-LgBiT (VTMLG) cells ^3^ were cultured in DMEM containing 10% FBS. Plasmids encoding HIV-GagPol-HiBiT and SARS-CoV-2 spike were described previously ^3^. Spike mutants were generated by standard mutagenesis procedures. hiVLP was produced by transient transfection of HEK293 cells with pHIV-GagPol-HiBiT and pSARS2-Spike-FLAG at a ratio of 1:1. Culture supernatants containing hiVLPs were collected 48 hours after transfection and filtered through a 0.45 µm Millex-HV filter (Merck).

### Modified hiVNT

VeroE6/TMPRSS2-LgBiT cells seeded in 96-well plates were washed and inoculated with hiVLP stocks (50 µl) containing diluted serum (1:20). At 3 hours after inoculation, cells were washed with PBS and treated with 100 µl of PBS containing DrkBiT peptide (1 µM) for 2 minutes. Cells were then added with 25 µl of 1x Nano-Glo Live Cell Substrate (Promega). Luciferase activity is measured with GloMax Discover System (Promega).

The luminescence signal inhibition (%) was calculated as follows:

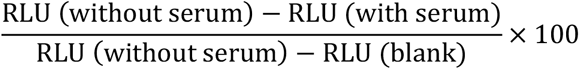

### Statistical analysis

The statistical significance of differences between two groups was evaluated by two-tailed unpaired t-test in the Prism 8 software (GraphPad).

## Data Availability

The datasets presented in this article are not readily available because it is difficult to ensure the de-identification of data. However, they can be available from the corresponding author on reasonable request. Requests to access the datasets should be directed to AR.

## Conflict of interests

YY is a current employee of Kanto Chemical Co. Inc. The authors have no conflicts of interest directly relevant to the content of this article.

## Acknowledgement

This study was supported by Rapid research and development Projects on COVID-19 of AMED (JP19fk0108110) to AR. We thank Kenji Yoshihara, Kazuo Horikawa, and Natsumi Takaira for their technical assistance. We acknowledge all patients and medical staff involved in the study.

## Notes

### Clinical Trial

N/A

### Author Declarations

This study was approved by Yokohama City University Certified Institutional Review Board (Reference No. B160800009, B200600115, B210300001), and the protocols used in the study were approved by the ethics committee.

